# Battling the COVID-19 Pandemic: Is Bangladesh Prepared?

**DOI:** 10.1101/2020.04.29.20084236

**Authors:** Md Hasinur Rahaman Khan, Tamanna Howlader, Md. Mazharul Islam

## Abstract

Following detection of the first few COVID-19 cases in early March, Bangladesh has stepped up its efforts to strengthen capacity of the healthcare system to avert a crisis in the event of a surge in the number of cases. This paper sheds light on the preparedness of the healthcare system by examining the spatial distribution of isolation beds across districts and divisions, forecasting the number of ICU units that may be required in the short term and analyzing the availability of frontline healthcare workers to combat the pandemic. As of May 2, COVID-19 cases have been found in 61 of the 64 districts in Bangladesh with Dhaka District being the epicenter. Seventy-one percent of the cases have been identified in 6 neighboring districts, namely, Dhaka, Narayanganj, Gazipur, Narsingdi, Munsiganj and Kishoreganj, which appear to form a spatial cluster. However, if one takes into account the population at risk, the prevalence appears to be highest in Dhaka, followed by Narayanganj, Gazipur, Kishorganj, Narsingdi and Munshiganj. These regions may therefore be flagged as the COVID-19 hotspots in Bangladesh. Among the eight divisions, prevalence is highest in Dhaka Division followed by Mymensingh. The number of cases per million exceeds the number of available isolation beds per million in the major hotspots indicating that there is a risk of the healthcare system becoming overwhelmed should the number of cases rise. This is especially true for Dhaka Division, where the ratio of COVID-19 patients to doctors appears to be alarmingly high. Mymensingh Division also has a disproportionately small number of doctors relative to the number of COVID-19 patients. Using second order polynomial regression, the analysis predicts that even if all ICU beds are allocated to COVID-19 patients, Bangladesh may run out of ICU beds soon after May 15, 2020. We conclude that in spite of a significant increase in hospital capacity during 2005-15 and a 57 % rise in the number of doctors during the same period, the healthcare system in Bangladesh and Dhaka Division in particular, may not be fully prepared to handle the COVID-19 crisis. Thus, further steps need to be taken to flatten the curve and improve healthcare capacity.

## 1 Introduction

Severe acute respiratory syndrome coronavirus (SARS-CoV-2), popularly known as COVID-19 is an infectious disease that was first identified in December 2019 in Wuhan. This is a respiratory infectious disease that has become a global pandemic within one and half months of its outbreak in Wuhan. Bangladesh found the first three coronavirus cases on 8 March 2020 that was confirmed by the Institute of Epidemilogy, Disease Control and Research (IEDCR) (IEDCR, 2020) at a press conference. The cases included two men and one woman, who were aged between 20 and 35 years. On March 16, the country detected three more cases of COVID-19 taking the total number of infected people to 8 (Khan & Hossain, 2020a). Bangladesh registered its first death due to the coronavirus on March 18. The country counted 8790 cases and 175 deaths till 2-nd of May (IEDCR, 2020). The fatality rate is 2.0 which is the second highest among South Asian countries and by this time over 76,063 tests have been performed. As of May 2 the situation is as follows: COVID-19 infections have already spread to 61 of the 64 districts; 6059 isolation beds are ready; 595 doctors, 546 nurses, 130 medical technologists and 350 other healthcare staff are allocated to treat COVID-19 patients (IEDCR, 2020). The burning question is, ‘Are these preparations adequate to fight the pandemic in Bangladesh?’

On March 7, when more that 100,000 people of 100 countries were affected by COVID-19, the World Health Organization (WHO) declared COVID-19 as a global pandemic (Shaw et al., 2020). Since then, majority of countries have imposed travel bans and lockdown of cities to stop the spread of COVID-19 and healthcare systems have taken unprecedented measures to expand their capacities to handle the surge. For instance, on 3rd February, China started sanitizing its city streets, public places, parks etc and later they introduced QR code for all residence to separate infected and non-infected people. Immediately after the outbreak, Wuhan city of China built two hospitals (30 ICU units and 1000 patients capacity) and several mobile cabins within a short period of time to meet excessive demand of hospital beds (Gao & Yu, 2020)(Christopher et al., n.d.). Meanwhile, the Ministry of Foreign Affairs, South Korea introduced government-supported quarantine facility for the returnees and the Central Disaster and Safety Countermeasures Headquarters implemented isolation, environmental disinfection, large scale diagnosis and epidemiological investigation. In order to prevent shortage of protective masks, the government introduced a countrywide five-day rationing system. Moreover, they hired 724 doctors amid the pandemic situation and allocated COVID-19 patients to infectious disease designated hospitals and separated 254 public hospitals for treating other diseases (Shaw et al., 2020).

COVID-19 pandemic has challenged global resource allocation and management system, especially in public health domain. The health professionals are threatened by the super-spreading behavior of novel Coronavirus; due to risk of infection while working in hospitals, large amount of protective equipment are in demand for both general people and health professionals. Although different countries have their own strategies to manage potential resources, merely few countries afford allocating necessary manpower and protective equipment at the right place in time. The current situation is new in many of the countries whereas some have previous experiences of dealing with SARS outbreak. On 20 January 2020, Taiwan announced its logistic support having 1100 negative-pressure isolation rooms, 1.9 million surgical and 44 million N95 masks respectively. The government implemented distribution of protective equipment through national security card which prevented panic buying of people [Wang et al. (2020), Shaw et al. (2020), Wang et al. (2020)]. Taiwan implemented its previous SARS epidemic experience to tackle the challenge by implementing modified Traffic Control Bundling (TCB) tool which provided small infection rate among health professionals during SARS outbreak in 2003 (Schwartz et al., 2020).

After mainland China, Europe has become the active center of COVID-19 - according to WHO’s declaration as of 13 March 2020 whereas the first case was reported in France on 24 January 2020 (Spiteri et al., 2020). About 47 laboratories in 31 of total 44 European countries had diagnostic capacity of COVID-19 by 31 January 2020 with a minimum facility of 8275 tests per week [Spiteri et al. (2020), Reusken et al. (2020)]. Germany made its first COVID-19 crisis response by closing border with neighboring countries and prohibiting the export of medical logistics including mask and other protective equipment (European Policy Institutes Network, 2020). The country has been expanding its intensive care facilities since January 2020, from 28000 intensive care beds to 40000 intensive care beds by far (Bennhold, 2020). On the other hand, Italy has suffered due to lack of effective measure and immediate response to the emergency, supply of protective equipment and inadequate medical stuffs. On March 11, one fifth of its ICU beds (1028 of 5200) were filled with COVID-19 patients (Remuzzi & Remuzzi, 2020). The country has total 5256 American Health Association (AHA) registered hospitals running with 534964 stuffs where 2704 of these hospitals provided ICU services through 96596 ICU beds. The Society of Critical Care Medicine advocates hospitals to implement a tiered staffing strategy during COVID-19 pandemic (Halpern et al., 2020).

In recent days COVID-19 infection has been increasing in South Asian countries. Despite having limited resources, Bhutan has firm control so far by early-sealing of its borders and monitoring the status regularly (Diplomat Risk Intelligence, 2020). For Bangladesh, COVID-19 is a humanitarian crisis with a public health dimension (World Economic Forum, 2020). While there are important lessons to be learnt from China, Italy, UK, USA, Spain, France and other developed countries, it may not be possible to adopt many of their policies in Bangladesh due to scarcity of resources. Developed countries have been able to invest heavily in their healthcare systems and this has enabled them to respond effectively to the COVID-19 pandemic. China, for example, has a health expenditure per capita that is 10 times that of Bangladesh (World Economic Forum, 2020). With limited resources, expanding healthcare capacity remains a challenge for Bangladesh. In 1980 there were only 28 ICU beds in Dhaka city. Since then the number of ICU beds has gradually increased (Nafseen, 2018). There are about one hundred hospitals with ICU facilities in Bangladesh and 80% of them are located in Dhaka (“Message from president. Criticon Bangladesh 2018”, 2018). Hospitals in Bangladesh currently have a total of 1,169 ICU beds. Out of these, 432 are in government hospitals and only 110 are outside the capital Dhaka, and 737 are in the private hospitals (Khan & Hossain, 2020b). These numbers are reasonably low if considered against a population of 170 milion (“The Daily DhakaTribune”, March 21, 2020). According to (Kennedy & Pronovost, 2006), the total number of ICU beds in a hospital should be between 5% and 12% depending on the care given by the hospital. In 2017-18, the total number of beds in hospitals were 1,27,360. Out of these, 48,934 were in government hospitals and 78426 were in private hospitals [(Nafseen, 2018). According to (“The Daily DhakaTribune”, March 21, 2020), Bangladesh currently has a total 141,903 hospital beds or 0.84 beds per 1000 people. Whether these resources are sufficient to tackle the COVID-19 pandemic requires a more indepth analysis. This paper tries to answer this question by examining the capacity of the healthcare system in Bangladesh in relation to the COVID-19 pandemic.

## 2 Methodology

The data are collected from two difference sources in Bangladesh. Information on number of COVID-19 cases in different administrative areas (divisions and districts of Bangladesh), number of isolation beds, and number of health professionals (doctors, nurses, medical technologists and other healthcare staff) assigned for treating COVID-19 patients are collected from https://corona.gov.bd, which is the government’s official web portal for reporting COVID-19 related information to the public (Institute of Epidemiology and Disease Control and Research, 2020). The data on population size of different districts and divisions are taken from projected population for the year 2021 (Population projection of Bangladesh: Dynamics and trends 2011-2061, Bangladesh Bureau of Statistics (BBS) in collaboration with Institute of Statistical Research and Training (ISRT), University of Dhaka) Bangladesh Bureau of Statistics (BBS) and Institute of Statistics Research and Training (ISRT), University of Dhaka (2015). Moreover, all of the maps presented in this study are produced by “mapReasy”, an R package for producing administrative maps Islam et al. (2017). We produced a number of spatial graphs with the above R package. We also predicted the number of COVID-19 infected people for a short period of time using second order polynomial regression.

## 3 Analysis

Figure 1 (left panel) shows the spatial distribution of laboratory confirmed cases of COVID-19 according to district. Cases have been identified in 61 of the 64 districts in Bangladesh. The epicenter is Dhaka district with 4419 infected persons, which accounts for about 50.2% of all cases in the country up till May 2. The contagion has spread to all districts that share a border with the epicenter. However among these, Dhaka’s neighboring district to the east, Narayanganj, appears to be the hardest hit registering 987 cases. Four neighboring north eastern districts, namely, Gazipur, Narsingdi, Munsiganj and Kishoreganj, have also seen alarming numbers of cases. These six regions seem to form a spatial cluster that accounts for 70.6% of all cases in Bangladesh. Other emerging hotspots include the port city of Chattogram, which is a major business center and Munshiganj district, which shares borders with Dhaka and Narayanganj. It is important to note, however, that the above results do not take into account the population size of each district, and therefore may not reflect the true scenario regarding the levels of infection in the populations. Figure 1 (right panel) shows the spatial distribution of laboratory confirmed cases of COVID-19 per million across districts. Dhaka is the number one hotspot with 300-349 cases per million while Narayanganj is second with 250-299 cases per million. The third most affected area is Gazipur with 80-99 cases per million followed by Munshiganj which has 60-79 cases per million. In contrast to the previous graph, Figure 1 (right panel) indicates that Gazipur has a higher number of cases per million than Kishoreganj. Furthermore, Mymensing and Cumilla are now no longer among the emerging hotspots having only 20-29 and 10-19 cases per million respectively.

**Figure 1:**
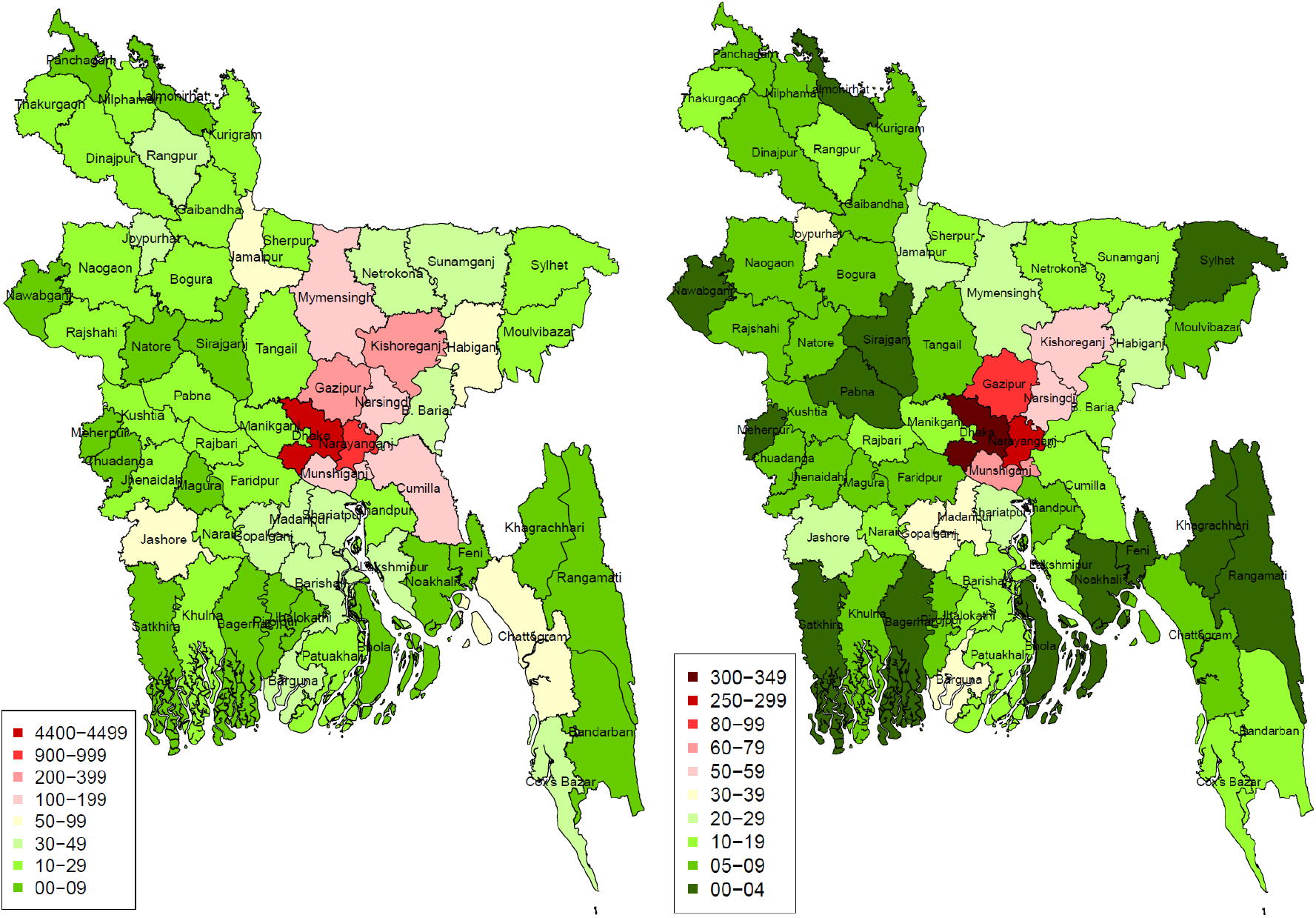
District-wise infected cases of COVID-19 in number (left panel) and in per million (right panel) in Bangladesh (as of May 2)

Bangladesh is divided into 8 administrative divisions. Figure 2 (left panel) shows the spatial distribution of laboratory confirmed cases of COVID-19 according to division. Dhaka Division is the epicenter with a total of 6415 cases up till May 2. The next highest number of cases have been reported in Chattogram Division, which is located to the south east of Dhaka Division followed by Mymensingh Division, which lies to the north. Both Rajshahi and Barisal Divisions, which lie in the far west and south of Bangladesh, respectively, less affected by the pandemic and have registered between 100 - 124 cases. The remaining three divisions are moderately affected and have managed to contain the number of cases within 125-149. However, to obtain a clearer picture regarding disease frequency in the divisions, one needs to take into account their population sizes as well. Figure 2 (right panel) shows the spatial distribution of laboratory confirmed cases of COVID-19 per million according to division. It is now seen that only Mymensingh have larger number of cases compared to Chattogram when adjusted for population size. This could be an indication of greater prevalence of COVID-19 in the former division relative to the latter.

**Figure 2:**
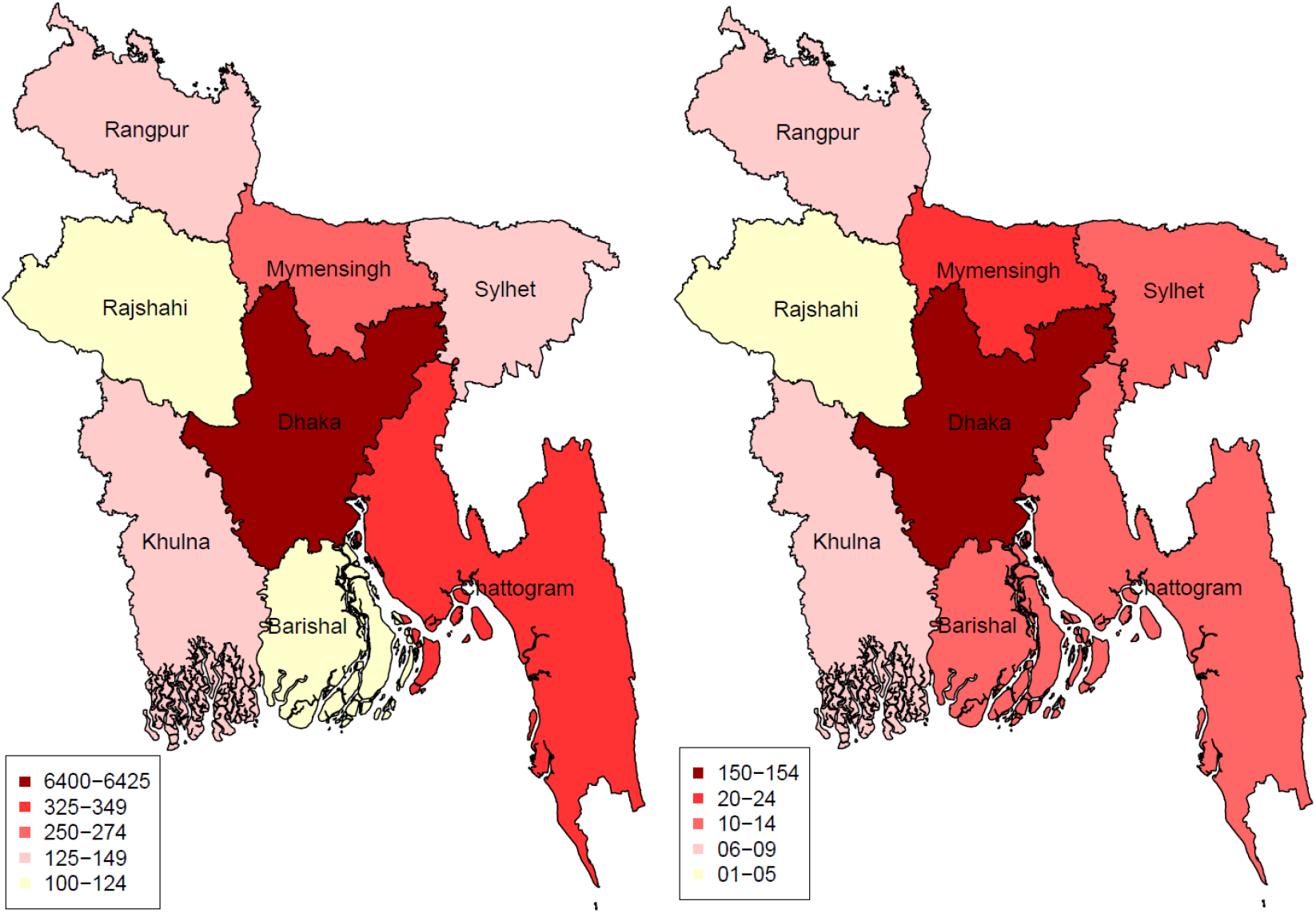
Division-wise infected cases of COVID-19 in number (left panel) and in per million (right panel) in Bangladesh (as of May 2)

The availability of sufficient number of isolation beds in affected areas is one indication of the preparedness of the health system in responding to a surge in the number of infected cases. Figure 3 (left panel) depicts the uneven distribution of number of isolation beds across districts in the country as of May 2. At a glance, there appears to be greater availability of isolation beds in districts to the north of Dhaka compared to the south. The epicenter Dhaka city has between 201-300 isolation beds. Among the northern districts, the number of available isolation beds are highest in Sherpur, Sylhet and Kishoreganj. Among the southern districts, Barishal, Chattogram and Rangamati possess the highest number of isolation beds, i.e. between 201-300, followed by Bhola, which has 176-200 isolation beds. The distribution of isolation beds suggests that some of the districts with high prevalence of confirmed cases could be at a risk of experiencing shortages should the number of cases rise. For instance, Naranganj district, which is the second major hotspot after Dhaka district with 4419 cases has between 126-150 isolation beds. Other COVID-19 hotspots, namely, Gazipur and Narsingdi, which have already registered 322 and 151 cases, respectively, have less than 50 isolation beds. Emerging hotspots such as Munshiganj having 122 cases, Madaripur and Gopalganj having between 40-49 confirmed cases also have less than 50 isolation beds. In contrast, health system facilities in districts such as Bhola, Cox’s Bazar, Naogaon, Joypurhat and Bogura are less likely to be overwhelmed as the number of cases (< 20) is far less than the available number of isolation beds (between 151 – 200). However, this is true only as long as the number of cases remains fairly low.

**Figure 3:**
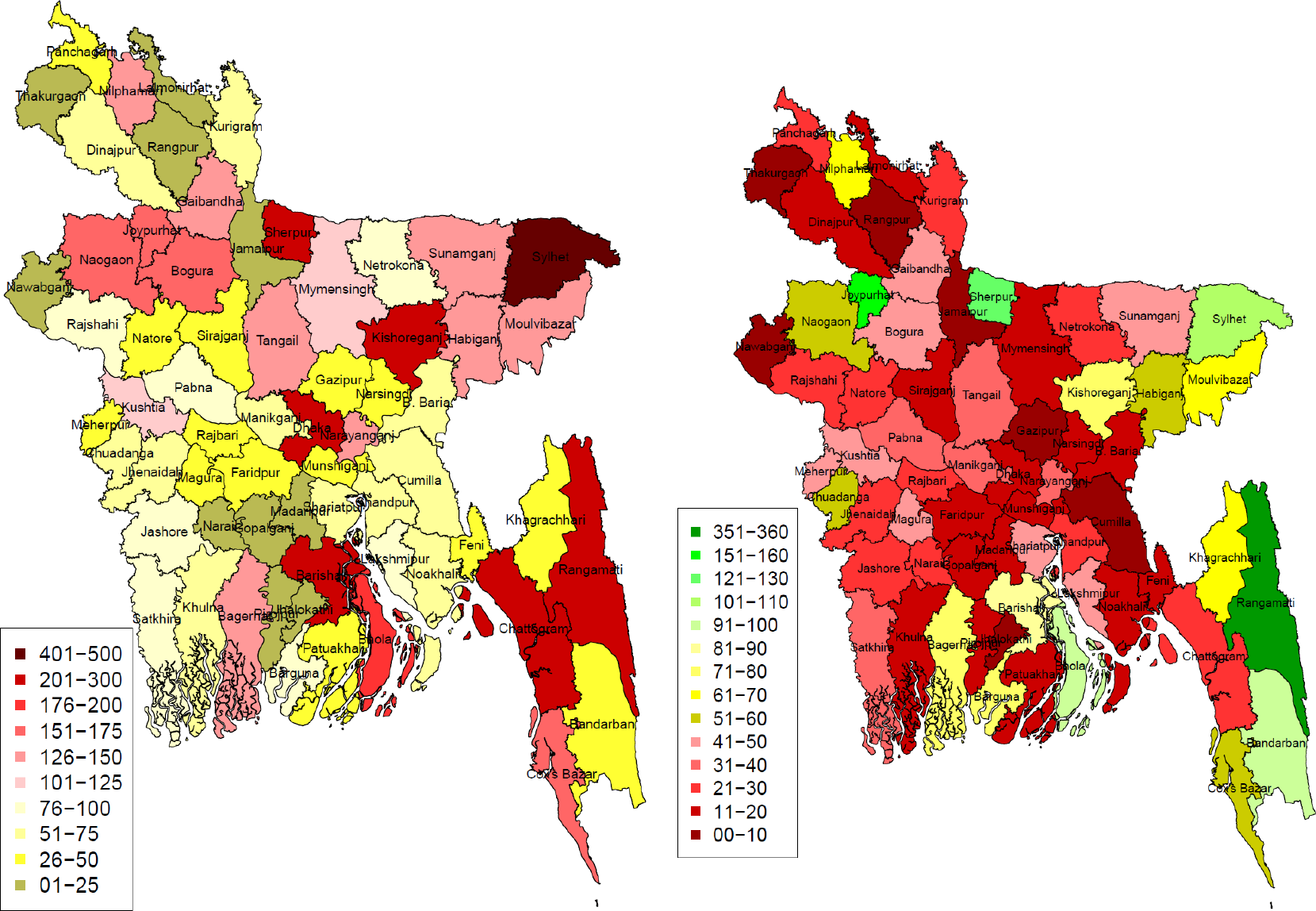
District-wise number of isolation beds in number (left panel) and in per million (right panel) in Bangladesh (as of May 2)

Figure 3 (right panel) provides further insights into health service capacity of Bangladesh by showing the number of isolation beds per million of the population in each of the 64 districts. Seventy-five percent of the districts have less than 50 isolation beds per million of the population. The epicenter, Dhaka, has between 11-20 isolation beds for every one million persons. Among the COVID-19 hotspots, Narayanganj has 31-40 isolation beds per million while Gazipur has less than 10. Chattogram has 21-30 isolation beds per million while Kishoreganj and Narsingdi have between 71-80 and 11-20 beds per million, respectively. It appears that Gazipur, Dhaka and Narsingdi districts are in a precarious situation due to the small number of isolation beds relative to the size of their populations. On the other hand, Rangamati, Bandarban and Bhola in the south, and Sherpur, Joypurhat and Sylhet in the north, have larger number of isolation beds relative to the sizes of their populations.

Figure 4 (left panel) shows the distribution of isolation beds with respect to Division. The largest number of isolation beds are available in the divisions of Dhaka and Chattogram. Chattogram Division has 1101-1200 beds while Dhaka Division has 1001-1100 beds. The third largest number of isolation beds is found in Sylhet Division with 901-1000 beds while Rajshahi and Khulna divisions have between 801-900 beds. Barishal, Mymensingh and Rangpur divisions have the lowest number of isolation beds. Although Dhaka and Chattogram appear to be better equipped relative to the other divisions based on this graph, the situation is reversed when population size is taken into account. Figure 4 (right panel) shows the number of isolation beds per million of the population in each of the eight divisions. Interestingly, Dhaka Division now has one of the lowest number of isolation beds when population size is taken into account with only 20-25 beds for every one million persons. Chattogram Division, which had the largest number of isolation beds according to Figure 4 (left panel), now falls behind Sylhet, Rajshahi, Khulna and Barishal with only 31-35 beds per million. Sylhet Division appears to be in the least vulnerable position with the highest number of isolation beds per million of its population.

**Figure 4:**
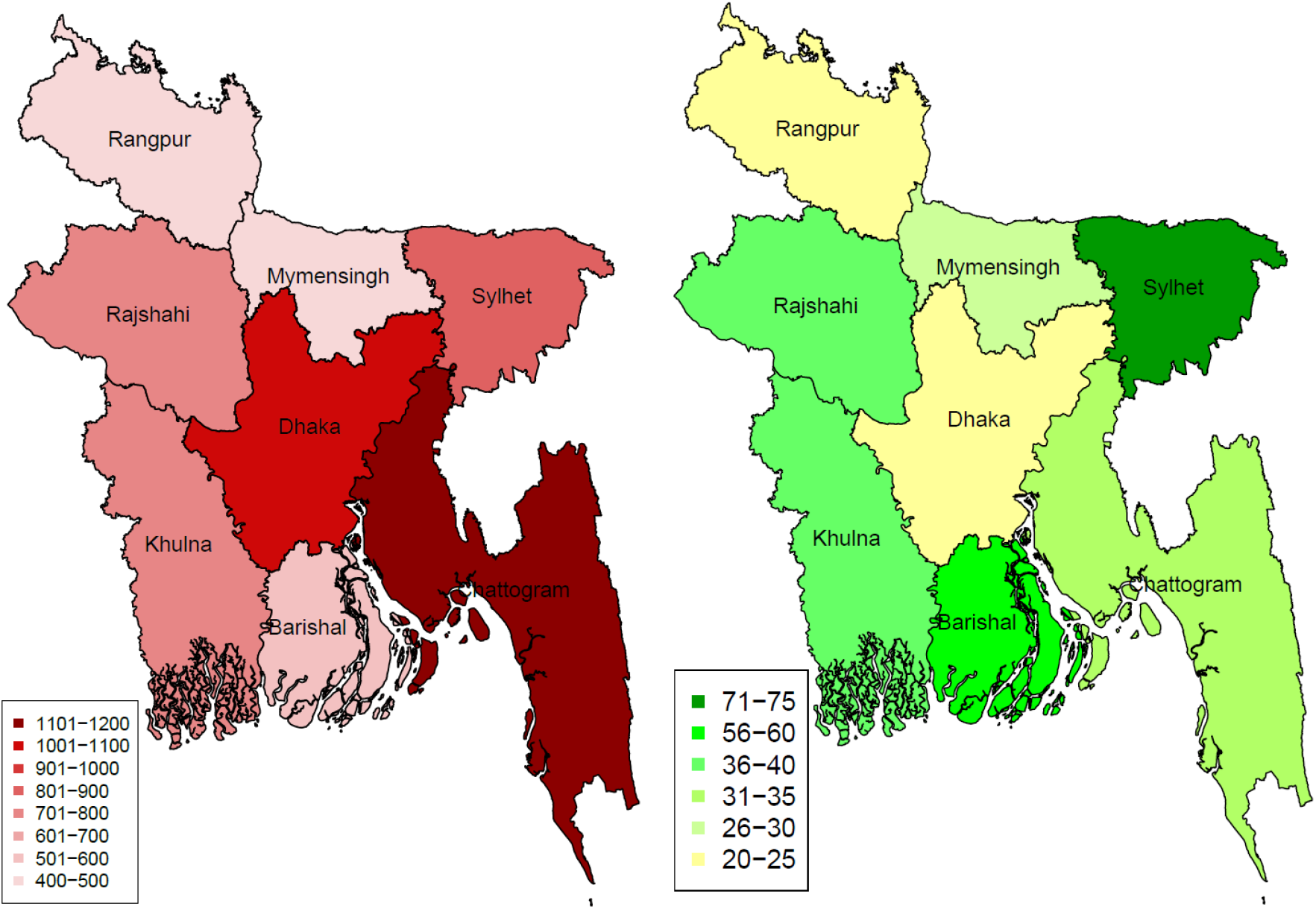
Division-wise number of isolation beds in number (left panel) and in per million (right panel) in Bangladesh (as of May 2)

The availability of qualified heath care workers is vital when it comes to combatting an epidemic. Figure 5 (left panel) summarizes the proportions of different types of healthcare workers in each division. In amost all the divisions, doctors form the largest proportion of healthcare staff followed by nurses. An exception is Rajshahi Division where the proportion of nurses exceeds the proportion of doctors. Sylhet and Khulna seem to have nearly equal proportions of doctors and nurses. The proportion of medical technologists appears to be highest in Mymensingh and lowest in Sylhet and Rajshahi. Apart from doctors, nurses and medical technologists, there are other supporting medical staff, such as ward boys, cleaners, etc who have vital roles within the health care system. A significant proportion of healthcare workers belong to this group in all the divisions. This is particularly true in Rajshahi Division where the proportion of health care workers belonging to the ‘Others’ category exceeds the proportion of doctors.

**Figure 5:**
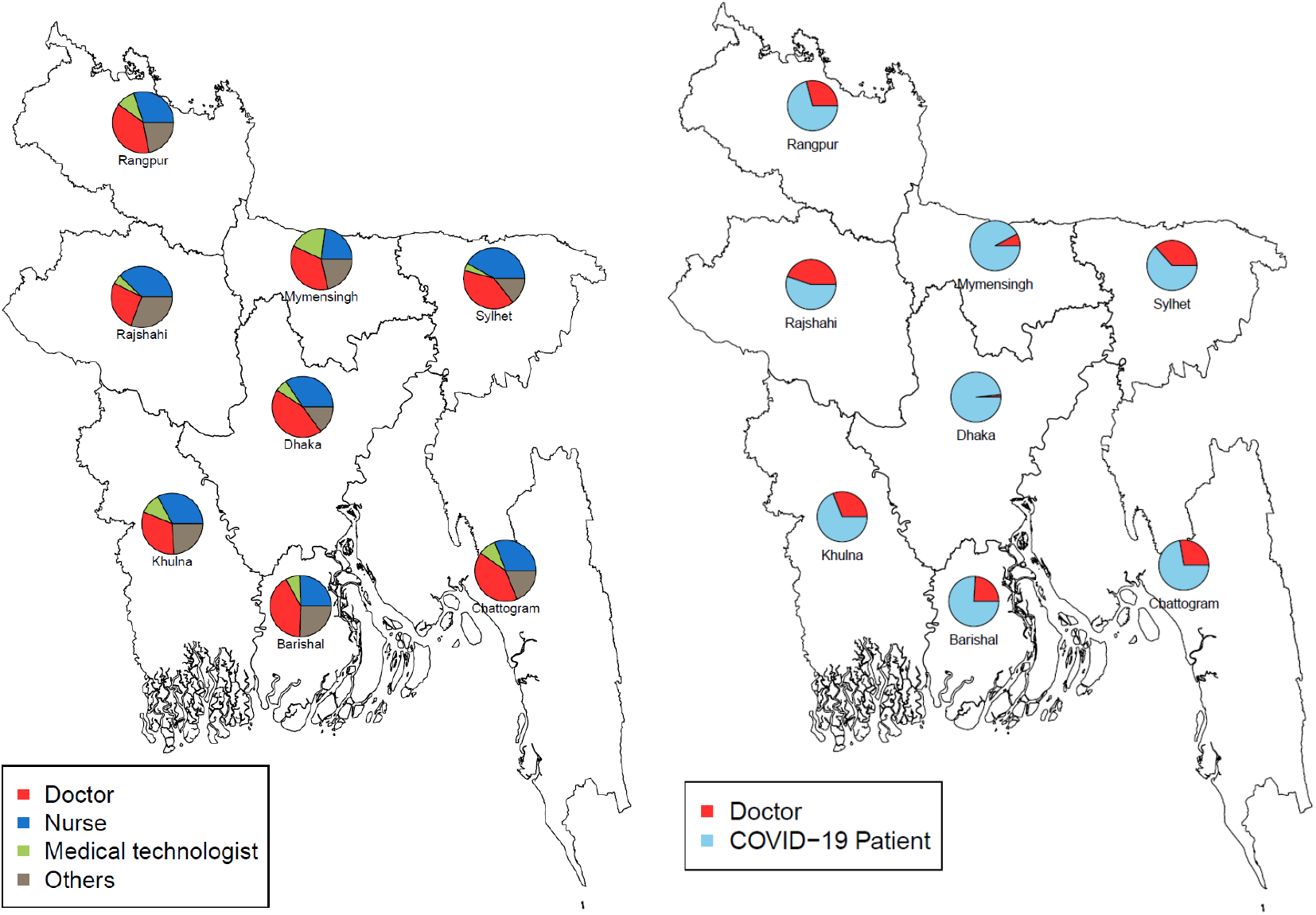
Division-wise distribution of health staff (left panel) and ratio of COVID-19 patients and doctors in the divisions of Bangladesh (right panel)

Figure 5 (right panel) shows the ratio of number of COVID-19 patients to the number of doctors available to treat them for each division according to data upto May 2. One alarming finding is the disproportionately small number of doctors in relation to the number of cases in Dhaka Division resulting in a very high ratio. The implication is a high risk of the healthcare system being overwhelmed due to acute shortage of doctors, who form the frontline defence in the fight against the pandemic. It appears that Mymensingh Division may also be in a vulnerable position due to the number of doctors being nearly 10% of the number of COVID-19 patients. Barisal Division, which has the lowest number of cases, appears to have nearly a quarter but the ratio could easily increase if the number of cases rise and more doctors cannot be provided to treat these cases. A similar situation exists in Chittagong Division as well. On the otherhand, Sylhet, Khulna, and Rangpur have smaller ratios and therefore appear to be in a better position to handle the crisis. Rajshahi division is in the best position to handle the crisis.

Figure 6 describes how the capacity of the healthcare system in Bangladesh has been evolving over the years. The figure shows two curves: the red curve for the number of hospital beds per 1000 people and the black curve for the number of doctors per 1000 people for the period 1970-2015 (WB, 2020). For some of the years, the ratios have been estimated through interpolation method before drawing this figure. It is seen that there was an increasing trend in the number of hospital beds per 1000 people from 1970 to 1991 during which the ratio increased from 0.1551 to 0.3081. Over the next 11 years the ratio remained relatively steady and then increased sharply after 2005 reaching a value of 0.8 per 1000 people in 2015. Thus, there has been a 166.6% increase in the number of hospital beds per 1000 people during 2005-2015 indicating significant improvements in healthcare infrastructure in the country (Khan & Hossain, 2020b). Figure 6 shows that there has been a slow exponential growth in the number of doctors per 1000 people during the 45 year period. Both curves are seen to converge in 2005 during which the number of hospital beds per 1000 people and the number of doctors per 1000 people were equal. Both ratios were equal to 0.3 meaning that for every 10,000 individuals in the population, there were 3 hospital beds and 3 doctors. Over the next ten years, the number of doctors per 1000 people increased by about 57% to a value of 0.472 in 2015. Overall, there has been growth in the healthcare sector but in a densely populated and developing country like Bangladesh, this growth may not be sufficient to fight a global pandemic.

**Figure 6:**
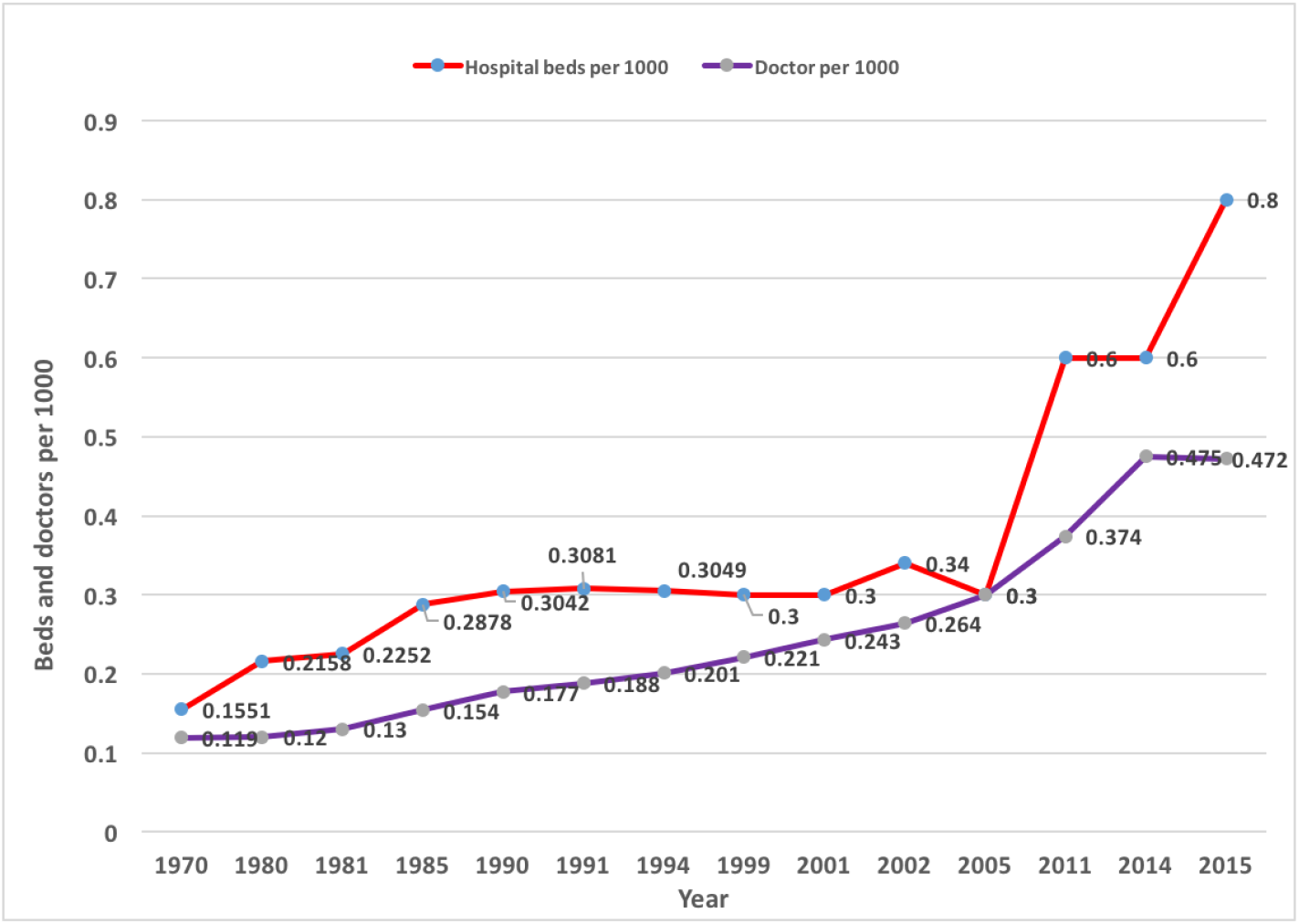
Yearly number of doctors and hospital beds per 1000 in Bangladesh

We analyzed the COVID-19 data of Bangladesh for predicting the number of infected cases by following the second order polynomial regression as suggested by (Khan & Hossain, 2020b), (Khan, 2020). Polynomial regression has been used in forcasting COVID-19 diseases along with for fitting trends by many researches [Pandeya et al. (2020), Johannes (2008), Howard (1943)]. During an epidemic, projections on the number of patients who may require intensive care are very useful to hospital administrators for planning. We therefore use the data till May 5 to obtain projected number of ICU patients in Bangladesh. According to (Khan & Hossain, 2020b), this projection is suitable for a short term period and hence projections were made for the period May 6 to June 12, 2020 based on the data reported from April 1 to May 5. Table 1 shows the projected numbers of infected people, hospital admissions, and ICU patients along with their associated 95% confidence intervals. The number of hospital admissions are estimated based on the findings in [Phua et al. (2020)] that 20% of infected patients require hospital admissions. Indirectly, this gives us estimates for the required number of isolation beds assuming an admitted person requires a single bed. Furthermore, according to (Phua et al., 2020), 6% of all reported cases need ICU admissions. This result was used to obtain the projected numbers of ICU patients reported in Table 1. Since each ICU patient requires a single bed, this table gives us, indirectly, projections for the required number of hospital beds in the ICU over the period May 6 to June 12, 2020.

**Table 1:** Projected total infections, hospital patients and ICU patients of COVID-19 in Bangladesh by polynomial regression method

| Date | Projected total infections | 95% CI for total infections |  | Projected total hospital patients | 95% CI for total hospital patients |  | Projected total ICU patients | 95% CI for total ICU patients |  |
| --- | --- | --- | --- | --- | --- | --- | --- | --- | --- |
|  |  | Lower | Upper |  | Lower | Upper |  | Lower | Upper |
| 6-May | 11595 | 11415 | 11774 | 2319 | 2283 | 2355 | 696 | 685 | 706 |
| 7-May | 12333 | 12150 | 12517 | 2467 | 2430 | 2503 | 740 | 729 | 751 |
| 8-May | 13095 | 12906 | 13285 | 2619 | 2581 | 2657 | 786 | 774 | 797 |
| 9-May | 13880 | 13684 | 14076 | 2776 | 2737 | 2815 | 833 | 821 | 845 |
| 10-May | 14687 | 14484 | 14891 | 2937 | 2897 | 2978 | 881 | 869 | 893 |
| 11-May | 15518 | 15306 | 15729 | 3104 | 3061 | 3146 | 931 | 918 | 944 |
| 12-May | 16371 | 16150 | 16592 | 3274 | 3230 | 3318 | 982 | 969 | 996 |
| 13-May | 17247 | 17016 | 17478 | 3449 | 3403 | 3496 | 1035 | 1021 | 1049 |
| 14-May | 18145 | 17904 | 18387 | 3629 | 3581 | 3677 | 1089 | 1074 | 1103 |
| 15-May | 19067 | 18813 | 19321 | 3813 | 3763 | 3864 | 1144 | 1129 | 1159 |
| 16-May | 20011 | 19745 | 20278 | 4002 | 3949 | 4056 | 1201 | 1185 | 1217 |
| 17-May | 20979 | 20699 | 21258 | 4196 | 4140 | 4252 | 1259 | 1242 | 1275 |
| 18-May | 21969 | 21675 | 22263 | 4394 | 4335 | 4453 | 1318 | 1301 | 1336 |
| 19-May | 22982 | 22673 | 23291 | 4596 | 4535 | 4658 | 1379 | 1360 | 1397 |
| 20-May | 24018 | 23693 | 24342 | 4804 | 4739 | 4868 | 1441 | 1422 | 1461 |
| 21-May | 25076 | 24735 | 25418 | 5015 | 4947 | 5084 | 1505 | 1484 | 1525 |
| 22-May | 26158 | 25799 | 26517 | 5232 | 5160 | 5303 | 1569 | 1548 | 1591 |
| 23-May | 27262 | 26885 | 27639 | 5452 | 5377 | 5528 | 1636 | 1613 | 1658 |
| 24-May | 28389 | 27994 | 28785 | 5678 | 5599 | 5757 | 1703 | 1680 | 1727 |
| 25-May | 29539 | 29124 | 29955 | 5908 | 5825 | 5991 | 1772 | 1747 | 1797 |
| 26-May | 30712 | 30277 | 31148 | 6142 | 6055 | 6230 | 1843 | 1817 | 1869 |
| 27-May | 31908 | 31452 | 32365 | 6382 | 6290 | 6473 | 1914 | 1887 | 1942 |
| 28-May | 33127 | 32648 | 33605 | 6625 | 6530 | 6721 | 1988 | 1959 | 2016 |
| 29-May | 34368 | 33867 | 34869 | 6874 | 6773 | 6974 | 2062 | 2032 | 2092 |
| 30-May | 35632 | 35109 | 36156 | 7126 | 7022 | 7231 | 2138 | 2107 | 2169 |
| 31-May | 36919 | 36372 | 37467 | 7384 | 7274 | 7493 | 2215 | 2182 | 2248 |
| 1-Jun | 38229 | 37658 | 38801 | 7646 | 7532 | 7760 | 2294 | 2259 | 2328 |
| 2-Jun | 39562 | 38965 | 40159 | 7912 | 7793 | 8032 | 2374 | 2338 | 2410 |
| 3-Jun | 40918 | 40295 | 41540 | 8184 | 8059 | 8308 | 2455 | 2418 | 2492 |
| 4-Jun | 42296 | 41647 | 42945 | 8459 | 8329 | 8589 | 2538 | 2499 | 2577 |
| 5-Jun | 43697 | 43022 | 44373 | 8739 | 8604 | 8875 | 2622 | 2581 | 2662 |
| 6-Jun | 45121 | 44418 | 45825 | 9024 | 8884 | 9165 | 2707 | 2665 | 2750 |
| 7-Jun | 46568 | 45837 | 47300 | 9314 | 9167 | 9460 | 2794 | 2750 | 2838 |
| 8-Jun | 48038 | 47278 | 50321 | 9608 | 9456 | 10064 | 2882 | 2837 | 3019 |
| 9-Jun | 49531 | 48741 | 50321 | 9906 | 9748 | 10064 | 2972 | 2924 | 3019 |
| 10-Jun | 51046 | 50226 | 51866 | 10209 | 10045 | 10373 | 3063 | 3014 | 3112 |
| 11-Jun | 52585 | 51734 | 53435 | 10517 | 10347 | 10687 | 3155 | 3104 | 3206 |
| 12-Jun | 54146 | 53264 | 55028 | 10829 | 10653 | 11006 | 3249 | 3196 | 3302 |

As discussed in (Khan & Hossain, 2020b), hospitals in Bangladesh have altogether 1,169 ICU beds in the year 2020. Most of the ICU beds are in the capital Dhaka. Government hospitals have nearly 40% of these beds i.e. 432 while the rest 737 are in the private hospitals. These numbers are reasonably low compared to the projected number of ICU beds irrespective of the division-wise distributions. According to Table 1, if all 1169 ICU beds are used for COVID-19 patients then Bangladesh will run out of ICU beds soon after May 15, 2020 and by end May there willl be cent percentage deficit of ICU beds. These findings sound the alarm that the healthcare system may not have the capacity to handle critical Covid-19 patients due to insufficient number of ICU beds.

## 4 Discussions and Conclusions

Following detection of the first few COVID-19 cases in early March, Bangladesh has stepped up its efforts to strengthen the capacity of the healthcare system to avert a crisis in the event of a surge in the number of cases. This analysis sheds light on the preparedness of the healthcare system in terms of the spatial distribution of number of isolation beds, availability of ICU beds and the availability of frontline healthcare workers to combat the pandemic. COVID-19 cases have been found in 61 of the 64 districts in Bangladesh as of May 2. Seventy-one percent of the cases have been identified in 6 neighboring districts that appear to form a spatial cluster of COVID-19 cases. These regions include Dhaka, Narayanganj, Gazipur, Narsingdi, Munsiganj and Kishoreganj with Dhaka being the epicenter. However if one takes into account the population at risk, the prevalence appears to be highest in Dhaka followed by Narayanganj, Gazipur, Kishorganj, Narsingdi, and Munshiganj. These regions may therefore be flagged as the COVID-19 hotspots in Bangladesh. Among the eight divisions, prevalence is highest in Dhaka Division followed by Mymensingh. Yet Dhaka Division has the lowest number of isolation beds per million of its population. A similar situation is seen in Mymensingh Division which also has high prevalence of the disease but a comparatively small number of isolation beds per million.

On a finer resolution, district-wise comparisons reveal that the epicenter, Dhaka District, with 300-349 cases per million has one of the lowest, i.e. between 11-20, isolation beds per million. Narajanganj, which is the major hotspot with 250-299 cases per million has only 31-40 isolation beds per million. The third hotspot is Gazipur with 80-99 cases per million and less than 10 isolation beds per million, which is the lowest among all the districts. Munsiganj, which is the fourth largest hotspot also has a small number of isolation beds relative to the number of cases per million of its population. These figures indicate that there is an elevated risk of the healthcare system becoming overwhelmed in the major hotspots in Bangladesh. On the positive side, the hilly district Rangamati has the highest number of isolation beds and zero cases per million in Bangladesh. With regard to availability of healthcare resource persons, the analysis finds that doctors form the largest proportion of healthcare staff in Bangladesh in five of the eight divisions. The proportions of doctors and nurses are nearly equal in Khulna and Sylhet Divisions. Rajshahi Division appears to be an outlier with the proportion of doctors being less than both the proportion of nurses and the proportion of other healthcare staff. An alarming finding is the very high ratio of COVID-19 patients to doctors in Dhaka Division. Mymensingh Division also has a disproportionately small number of doctors available to treat COVID-19 patients.

### 4.1 Recommendations and policy implications

The analysis has revealed that the numbers of isolation beds and doctors available to treat COVID-19 patients are worryingly low in the major hotspots in Bangladesh. This finding has important implications for policy. In a densely populated country like Bangladesh where majority of the people are poor, it is difficult to effectively enforce social distancing and other preventive measures. Thus, there is a fear that the number of cases could flare up at any time during the course of the pandemic. In these circumstances, the healthcare system must be adequately prepared to face the looming crisis by creating excess capacity and mobilizing resources to the most affected areas.

Based on the analysis, we recommend that the number of isolation beds in Dhaka District (which includes Dhaka city) as well as Gazipur District be more than tripled so as to be on par with the number of cases. In Narayanganj District the number of isolation beds should at least be doubled. Projections indicate that there is a risk of a deficit in the number of ICU beds for critical patients. Thus, increasing the number of ICU units should also be a priority. Strategies for increasing capacity could include freeing up more beds for COVID-19 patients in hospitals, increasing the number of COVID-19 designated healthcare facilities by roping in private hospitals and clinics, and repurposing government buildings and setting up camps as an emergency measure should the outbreak go out of hand. With the number of doctors available to treat COVID-19 patients being dangerously low in Dhaka Division, we recommend increasing the number of doctors by several folds. This could be done by recruiting new doctors in hospitals including final year medical students and calling retired doctors back to work. There have been reports that some doctors avoid work due to fear of contracting the virus due to lack of PPE. As a result, both COVID-19 patients as well as patients with other medical conditions may not receive adequate medical care. Lack of PPE may therefore be identified as one of the leading causes of shortages in medical personnel.

We recommend that in addition to offering attractive incentives to frontline healthcare workers, the government must by all means ensure that medical personnel on duty receive PPE and training on how to use them. This is one of several measures that need to be taken to ensure the safety of healthcare workers and their families so as to maximize their participation in the fight against the pandemic. COVID-19 preparedness in Bangladesh must include an effective and feasible plan of action in which guidance on resource management and communication and coordination between national health stakeholders are of utmost importance. If preparations are taken in a pragmatic way, Bangladesh will win the COVID-19 war with less fatalities since it has a strong network of community health workers, a history of success in public-private partnerships during emergencies like floods and cyclones, and people with incredible levels of resilience.

## Data Availability

The working data set used for this study has been submitted to the journal as additional supporting file.

## Author’s Contributions

MHRK contributed to designing, analysing and drafting the manuscript, TH contributed to analyse and finalized the drafting, MMI contributed to arranging and analysing the data.

